# Prognostic Significance of Cerebrospinal Fluid Glucose, Protein, and White Blood Cell Count in Breast Cancer Leptomeningeal Disease

**DOI:** 10.64898/2026.02.07.26345775

**Authors:** Sugam Gouli, Sujan Niraula, Andrea Baran, Huina Zhang, Ruth O’Regan, Nimish Mohile, Carey K Anders, Sara Hardy, Ajay Dhakal

## Abstract

**Background:** Leptomeningeal disease (LMD) is a serious complication of metastatic breast cancer (MBC) with poor survival. This single-institution retrospective study compares overall survival (OS) among MBC patients with LMD based on CSF parameters (glucose, protein, and WBC count)

**Methodology:** MBC patients who were diagnosed with LMD between 2010-2023 at Wilmot Cancer Institute were screened for eligibility. Only those with available data on CSF glucose, protein, and WBC count were included. OS was assessed via the Kaplan-Meier method and compared using the log-rank test. Cox models were used for multivariate analysis.

**Results:** Out of 69 patients with MBC LMD, 28 had CSF data and were included in the final analysis. The CSF cytology-positive cohort had significantly lower glucose levels vs the CSF cytology-negative cohort [median (IQR) 40 (18-58) vs 64 (53-92) mg/dl, p=0.006]. Median CSF WBC count was significantly higher in the CSF cytology positive cohort vs the CSF cytology negative cohort [median (IQR) 13 (6-44) vs. 2(2-4)cells/mm^3^, p=0.001]. When stratified by CSF cytology results and CSF glucose levels, the CSF cytology negative, glucose-low group was associated with the worst OS, while the CSF cytology negative, normal/high glucose group was associated with the best OS(*p*=0.03) in an unadjusted analysis. Multivariate analysis confirmed that low CSF glucose was independently associated with poorer survival [HR 4.64 (1.71, 13.2)]. Neither CSF protein levels nor CSF WBC counts were significantly associated with OS in unadjusted and multivariate analyses.

**Conclusion:** Low CSF glucose was associated with worse OS than normal/high CSF glucose. There was insufficient evidence to suggest that CSF protein or CSF WBC counts were associated with OS.

## Introduction

Breast cancer is among the most common solid tumors that give rise to leptomeningeal disease (LMD). Ten to twenty percent of breast cancer patients who have parenchymal brain metastases eventually develop LMD.^1^ LMD is the spread of breast cancer to the leptomeninges and has high rates of morbidity and mortality. The median overall survival is 3-4 months; however, some patients survive for years, indicating variability in survival.^2^ Hence, it is crucial to have a clear understanding of various prognostic factors while predicting the survival of patients with breast cancer LMD. There is limited data on whether standard cerebrospinal fluid (CSF) parameters, such as glucose, protein, and white blood cell (WBC) counts, have prognostic significance.^2,3^ Our retrospective study investigates the prognostic significance of CSF protein, glucose, and WBC count in metastatic breast cancer with LMD.

## Material and Methods

### Design/Settings/Participants

Breast cancer patients diagnosed with LMD between January 1^st,^ 2010, and January 1^st^, 2023, who were treated at Wilmot Cancer Institute, University of Rochester Medical Center (URMC), were identified from the institutional database. The URMC Institutional Review Board (IRB) approved this retrospective study. LMD diagnosis was confirmed through a magnetic resonance imaging (MRI) report and/or a positive CSF cytology study. Patients aged 18 years or older, with imaging or cytology/pathology-proven LMD and medical records available for review, were eligible. Only eligible patients with available CSF glucose, protein, and WBC count data were included in the analyses.

### Variables

The study’s primary aim was to assess the association of CSF protein, glucose, and WBC count in LMD patients and overall survival (OS). Eligible patients were categorized into groups based on CSF glucose level (low vs normal or high), CSF protein level (high vs normal or low), and CSF WBC count (high vs normal). URMC’s standard CSF normal parameter ranges were used to define the levels of glucose (normal 50-80mg/dL), protein (normal 12-60mg/dL), and WBC counts (normal 0-5 cells/mm3) in the study.

### Statistical methods

Baseline characteristics were compared using Fisher’s Exact test for categorical variables and the Wilcoxon rank-sum test for continuous variables. Overall survival (OS) was defined as the time from the diagnosis of LMD to the date of death from any cause or date of last follow-up, if alive at the end of the study period. OS among patient subsets was graphically summarized using the Kaplan-Meier method and compared using the log-rank test. Multivariate Cox proportional-hazards regression models were used to estimate the risk of death associated with the subset while adjusting for other potential prognostic factors. A stepwise model selection process is used to select a parsimonious model from a list of potential covariates after including the subset of interest in the model. The list of variables included performance status, disease intervals, CSF cytology, and receptor subtypes (HER2+, ER+/HER2-, triple-negative breast cancer). Variables meeting entry criteria (*p* <.10) and staying criteria (*p* <.05) were retained in the final Cox model. All reported *p*-values are 2-sided, with *p* <.05 indicating statistical significance. All analyses were performed in SAS 9.4 (SAS Institute, Inc., Cary, NC, USA).

## Results

Out of 219 potential LMD patients screened, 69 patients had LMD with breast cancer. Of 69 patients, only 28 had CSF glucose, protein, and WBC data available and were included in the final analyses (Figure 1). The median age of the study cohort was 52 years; predominantly white (89.3%); 50.0% ECOG performance status 0-1; 7.1% de novo LMD; 39.3% TNBC; 21.4% HER2+; 39.3% ER+/HER2-. Patient and disease characteristics within each patient subset are shown in Table 1.

**Table 1.** Patient/Disease Characteristics by Sub-groups.

| Variables | CSF Glucose (mg/dl) |  |  | CSF Protein (mg/dl) |  |  | CSF WBC Count/mm<sup>3</sup> |  |  |
| --- | --- | --- | --- | --- | --- | --- | --- | --- | --- |
|  | Low (N=11) | normal/<br>high (N=17) | p-value | Normal/<br>Low (N=7) | High<br>(N=21) | p-value | Normal<br>(N=17) | High<br>(N=14) | p-value |
| **Age** (median (IQR)) | 48.0 (38.0-63.0) | 58.0 (48.0-64.0) | 0.22 | 52.0<br>(40.0-58.0) | 52.0<br>(48.0-69.0) | 0.36 | 52.0<br>(45.0-61.0) | 55.0<br>(48.0-63.0) | 0.69 |
| **Race** |  |  | 0.70 |  |  | 0.01 |  |  | 1.00 |
| White | 11 (100.0%) | 14 (82.4%) |  | 4 (57.1%) | 21 (100%) |  | 15<br>(88.2%) | 13<br>(92.9%) |  |
| Black | 0 | 2 (11.8%) |  | 2 (28.6%) | 0 |  | 1 (5.9%) | 1 (7.1%) |  |
| Other | 0 | 1 (5.9%) |  | 1 (14.3%) | 0 |  | 1 (5.9%) | 0 |  |
| **Sex** |  |  |  |  |  |  |  |  |  |
| Female | 11 (100%) | 17 (100%) |  | 7 (100%) | 21 (100%) |  | 17<br>(100%) | 14<br>(100%) |  |
| **Performance status** |  |  | 0.43 |  |  | 1.00 |  |  | 0.84 |
| KPS 70-100 or<br>ECOG 0-1 | 6 (54.6%) | 8 (47.1%) |  | 4 (57.1%) | 10<br>(47.6%) |  | 10<br>(58.8%) | 7<br>(50.0%) |  |
| KPS 50-70 or ECOG<br>2 | 4 (36.4%) | 9 (52.9%) |  | 3 (42.9%) | 10<br>(47.6%) |  | 6<br>(35.3%) | 7<br>(50.0%) |  |
| KPS<50 or ECOG >2 | 1 (9.1%) | 0 |  | 0 | 1 (4.8%) |  | 1 (5.9%) | 0 |  |
| **Interval from**<br>**primary breast**<br>**cancer to LMD**<br>**diagnosis** |  |  | 0.09 |  |  | 0.90 |  |  | 0.15 |
| De novo | 0 | 2 (11.8%) |  | 0 | 2 (9.5%) |  | 0 | 2 (14.3%) |  |
| <36 mo | 5 (45.5%) | 4 (23.5%) |  | 2 (28.6%) | 7 (33.3%) |  | 4 (23.5%) | 6 (42.9%) |  |
| 37-72 months | 2 (18.2%) | 0 |  | 0 | 2 (9.5%) |  | 1 (5.9%) | 1 (7.1%) |  |
| >72 mo | 4 (36.4%) | 11 (64.7%) |  | 5 (71.4%) | 10 (47.6%) |  | 12 (70.6%) | 5 (35.7%) |  |
| **Interval from metastatic disease to LMD diagnosis** |  |  | 0.61 |  |  | 0.60 |  |  | 0.046 |
| De novo | 3 (27.3%) | 3 (17.7%) |  | 1 (14.3%) | 5 (23.8%) |  | 1 (5.9%) | 5 (35.7%) |  |
| <12 mo | 2 (18.2%) | 5 (29.4%) |  | 3 (42.9%) | 4 (19.1%) |  | 3 (17.7%) | 4 (28.6%) |  |
| 12-60 months | 4 (36.4%) | 3 (17.7%) |  | 2 (28.6%) | 5 (23.8%) |  | 5 (29.4%) | 4 (28.6%) |  |
| >60 mo | 2 (18.2%) | 6 (35.3%) |  | 1 (14.3%) | 7 (33.3%) |  | 8 (47.1%) | 1 (7.1%) |  |
| **Interval from parenchymal brain metastasis to LMD diagnosis** |  |  | 1.00 |  |  | 0.84 |  |  | 0.46 |
| De novo | 5 (45.5%) | 7 (41.2%) |  | 3 (42.9%) | 9 (42.9%) |  | 8 (47.1%) | 5 (35.7%) |  |
| <12 mo | 1 (9.1%) | 3 (17.7%) |  | 1 (14.3%) | 3 (14.3%) |  | 3 (17.7%) | 2 (14.3%) |  |
| 12+months | 1 (9.1%) | 1 (5.9%) |  | 1 (14.3%) | 1 (4.8%) |  | 2 (11.8%) | 0 |  |
| no parenchymal disease | 4 (36.4%) | 6 (35.3%) |  | 2 (28.6%) | 8 (38.1%) |  | 4 (23.5%) | 7 (50.0%) |  |
| **Bone metastasis at LMD diagnosis** | 4 (36.4%) | 16 (94.1%) | 0.002 | 6 (85.7%) | 14 (66.7%) | 0.63 | 14 (82.4%) | 7 (50.0%) | 0.12 |
| **Lymph node metastasis at LMD diagnosis** | 11 (100.0%) | 12 (70.6%) | 0.12 | 6 (85.7%) | 17 (81.0%) | 1.00 | 13 (76.5%) | 13 (92.9%) | 0.34 |
| **Lung metastasis at LMD diagnosis** | 0 | 4 (23.5%) | 0.13 | 1 (14.3%) | 3 (14.3%) | 1.00 | 5 (29.4%) | 0 | 0.048 |
| **Liver metastasis at LMD diagnosis** | 1 (9.1%) | 9 (52.9%) | 0.04 | 4 (57.1%) | 6 (28.6%) | 0.21 | 8 (47.1%) | 2 (14.3%) | 0.07 |
| **ESMO LMD type** |  |  | 0.006 |  |  | 1.00 |  |  | 0.001 |
| I | 9 (81.8%) | 4 (23.5%) |  | 3 (42.9%) | 10 (47.6%) |  | 3 (17.7%) | 11 (78.6%) |  |
| II | 2 (18.2%) | 13 (76.5%) |  | 4 (57.1%) | 11 (52.4%) |  | 14 (82.4%) | 3 (21.4%) |  |
| **Anatomical Involvement of LMD** |  |  | 0.60 |  |  | 0.26 |  |  | 0.33 |
| Brain | 4 (36.4%) | 6 (35.3%) |  | 2 (28.6%) | 8 (38.1%) |  | 6 (35.3%) | 4 (28.6%) |  |
| Spinal cord | 1 (9.1%) | 5 (29.4%) |  | 2 (28.6%) | 4 (19.1%) |  | 5 (29.4%) | 3 (21.4%) |  |
| Both (the brain and the spinal cord) | 5 (45.5%) | 4 (23.5%) |  | 1 (14.3%) | 8 (38.1%) |  | 6 (35.3%) | 4 (28.6%) |  |
| NA (cytological involvement only) | 1 (9.1%) | 2 (11.8%) |  | 2 (28.6%) | 1 (4.8%) |  | 0 | 3 (21.4%) |  |
| **Number of sites of LMD involvement** |  |  | 0.91 |  |  | 0.19 |  |  | 0.21 |
| 1 | 1 (9.1%) | 4 (23.5%) |  | 2 (28.6%) | 3 (14.3%) |  | 3 (17.7%) | 3 (21.4%) |  |
| 2 | 1 (9.1%) | 1 (5.9%) |  | 0 | 2 (9.5%) |  | 1 (5.9%) | 1 (7.1%) |  |
| More than 2 | 8 (72.7%) | 10 (58.8%) |  | 3 (42.9%) | 15 (71.4%) |  | 13 (76.5%) | 7 (50.0%) |  |
| NA cytological only | 1 (9.1%) | 2 (11.8%) |  | 2 (28.6%) | 1 (4.8%) |  | 0 | 3 (21.4%) |  |
| **Parenchymal brain metastasis at the time of LMD diagnosis** |  |  | 0.66 |  |  | 1.00 |  |  | 0.10 |
| Present | 5 (45.5%) | 8 (47.1%) |  | 3 (42.9%) | 10 (47.6%) |  | 11 (64.7%) | 4 (28.6%) |  |
| Absent | 5 (45.5%) | 9 (52.9%) |  | 4 (57.1%) | 10 (47.6%) |  | 6 (35.3%) | 9 (64.3%) |  |
| Unknown | 1 (9.1%) | 0 |  | 0 | 1 (4.8%) |  | 0 | 1 (7.1%) |  |
| **Number of parenchymal brain metastasis 1 cm or larger** |  |  | 0.83 |  |  | 0.29 |  |  | 0.27 |
| 0 | 6 (54.5%) | 11 (64.7%) |  | 3 (42.9%) | 14 (66.7%) |  | 8 (47.1%) | 10 (71.4%) |  |
| 1-2 | 3 (27.3%) | 4 (23.5%) |  | 2 (28.6%) | 5 (23.8%) |  | 7 (41.2%) | 2 (14.3%) |  |
| More than 2 | 1 (9.1%) | 0 |  | 0 | 1 (4.8%) |  | 1 (5.9%) | 0 |  |
| Unknown | 1 (9.1%) | 2 (11.8%) |  | 2 (28.6%) | 1 (4.8%) |  | 1 (5.9%) | 2 (14.3%) |  |
| **Associated hydrocephalus** |  |  | 0.12 |  |  | 0.21 |  |  | 0.48 |
| Yes | 6 (54.6%) | 4 (23.5%) |  | 4 (57.1%) | 6 (28.6%) |  | 5 (29.4%) | 6 (42.9%) |  |
| No | 5 (45.5%) | 13 (76.5%) |  | 3 (42.9%) | 15 (71.4%) |  | 12 (70.6%) | 8 (57.1%) |  |
| **Symptomatic from LMD** |  |  | 1.00 |  |  | 1.00 |  |  | 1.00 |
| Yes | 11 (100%) | 16 (94.1%) |  | 7 (100%) | 20 (95.2%) |  | 16 (94.1%) | 14 (100%) |  |
| No | 0 | 1 (5.9%) |  | 0 | 1 (4.8%) |  | 1 (5.9%) | 0 |  |
| **Radiation treatment of LMD** | 9 (81.8%) | 11 (64.7%) | 0.42 | 5 (71.4%) | 15 (71.4%) | 1.00 | 13 (76.5%) | 10 (71.4%) | 1.00 |
| **Systemic therapy to treat LMD** | 5 (45.5%) | 16 (94.1%) | 0.007 | 7 (100%) | 14 (66.7%) | 0.14 | 14 (82.4%) | 9 (64.3%) | 0.41 |
| **Intrathecal therapy to treat LMD** | 0 | 1 (5.9%) | 1.00 | 0 | 1 (4.8%) | 1.00 | 0 | 1 (7.1%) | 0.45 |
| **No treatment for LMD** | 1 (9.1%) | 0 | 0.39 | 0 | 1 (4.8%) | 1.00 | 0 | 1 (7.1%) | 0.45 |
| **Radiological involvement (11 missing)** |  |  | 0.81 |  |  | 0.02 |  |  | 0.26 |
| Linear | 0 | 1 (9.1%) |  | 0 | 1 (8.3%) |  | 2 (18.2%) | 0 |  |
| Nodular | 4 (66.7%) | 8 (72.7%) |  | 2 (40.0%) | 10 (83.3%) |  | 8 (72.7%) | 5 (55.6%) |  |
| Both (linear and nodular) | 1 (16.7%) | 0 |  | 0 | 1 (8.3%) |  | 0 | 2 (22.2%) |  |
| Normal leptomeninges | 1 (16.7%) | 2 (18.2%) |  | 3 (60.0%) | 0 |  | 1 (9.1%) | 2 (22.2%) |  |
| **Breast cancer group** |  |  | 0.69 |  |  | 1.00 |  |  | 0.002 |
| ER+/HER2- | 3 (27.3%) | 8 (47.1%) |  | 3 (42.9%) | 8 (38.1%) |  | 10 (58.8%) | 2 (14.3%) |  |
| HER2+ | 3 (27.3%) | 3 (17.7%) |  | 1 (14.3%) | 5 (23.8%) |  | 5 (29.4%) | 2 (14.3%) |  |
| TNBC | 5 (45.5%) | 6 (35.3%) |  | 3 (42.9%) | 8 (38.1%) |  | 2 (11.8%) | 10 (71.4%) |  |

**Figure 1.**
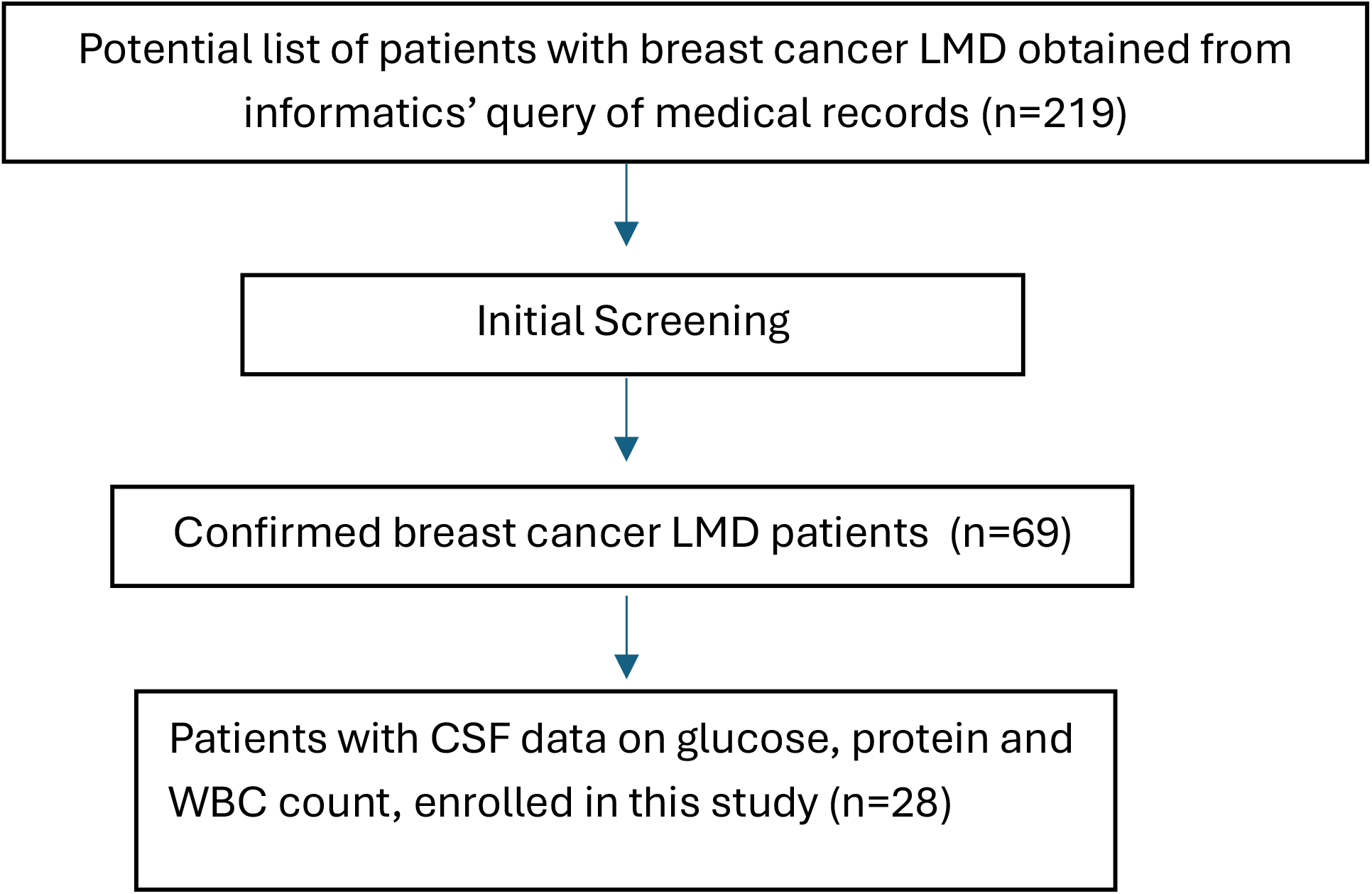
Flow Diagram Showing Patient Selection

Median CSF glucose, protein levels, and median CSF WBC cell counts were compared between the CSF cytology positive vs negative cohort (Table 2). The CSF cytology-positive cohort had significantly lower glucose levels than the CSF cytology-negative cohort [median (IQR) of 40 (18-58) vs. 64 (53-92) mg/dl, p=0.006]. Similarly, the median CSF WBC count was significantly higher in the CSF cytology positive cohort vs the CSF cytology negative cohort [median (IQR) 13 (6-44) vs. 2(2-4)cells/mm^3^, p=0.001]. There was no significant difference in the CSF protein level among the cohorts. A prior study from the same database showed that the CSF cytology-negative group had better OS than the CSF cytology-positive group.^4^ Hence, the CSF glucose and WBC count subsets were further stratified by CSF cytology status to eliminate potential confounding by cytology status when comparing OS between groups.

**Table 2.** CSF Parameters Based on CSF Cytology.

|  | CSF Cytology (median(IQR)) |  | p-value |
| --- | --- | --- | --- |
|  | Positive (N=13) | Negative (N=11) |  |
| CSF glucose mg/dl | 40(18-58) | 64(53-92) | 0.006 |
| CSF protein mg/dl | 99(59-234) | 74(46-185) | 0.68 |
| CSF WBC<br>count/mm<sup>3</sup> | 13(6-44) | 2(2-4) | 0.001 |

*Survival based on CSF Glucose:* In an unadjusted analysis, there was a significant difference in OS between CSF cytology positive/glucose low, CSF cytology positive/glucose normal or high, CSF cytology negative/glucose low, and CSF cytology negative/glucose normal or high groups (p=0.03; in Table 3, Figure 2a). CSF cytology negative/glucose normal or high group had the longest OS, and CSF cytology negative, glucose low had the shortest OS. Low CSF glucose was associated with shorter OS in multivariate analysis (Table 4) [adjusted hazard ratio (HR) 4.64 (1.71, 13.2)].

**Table 3.**
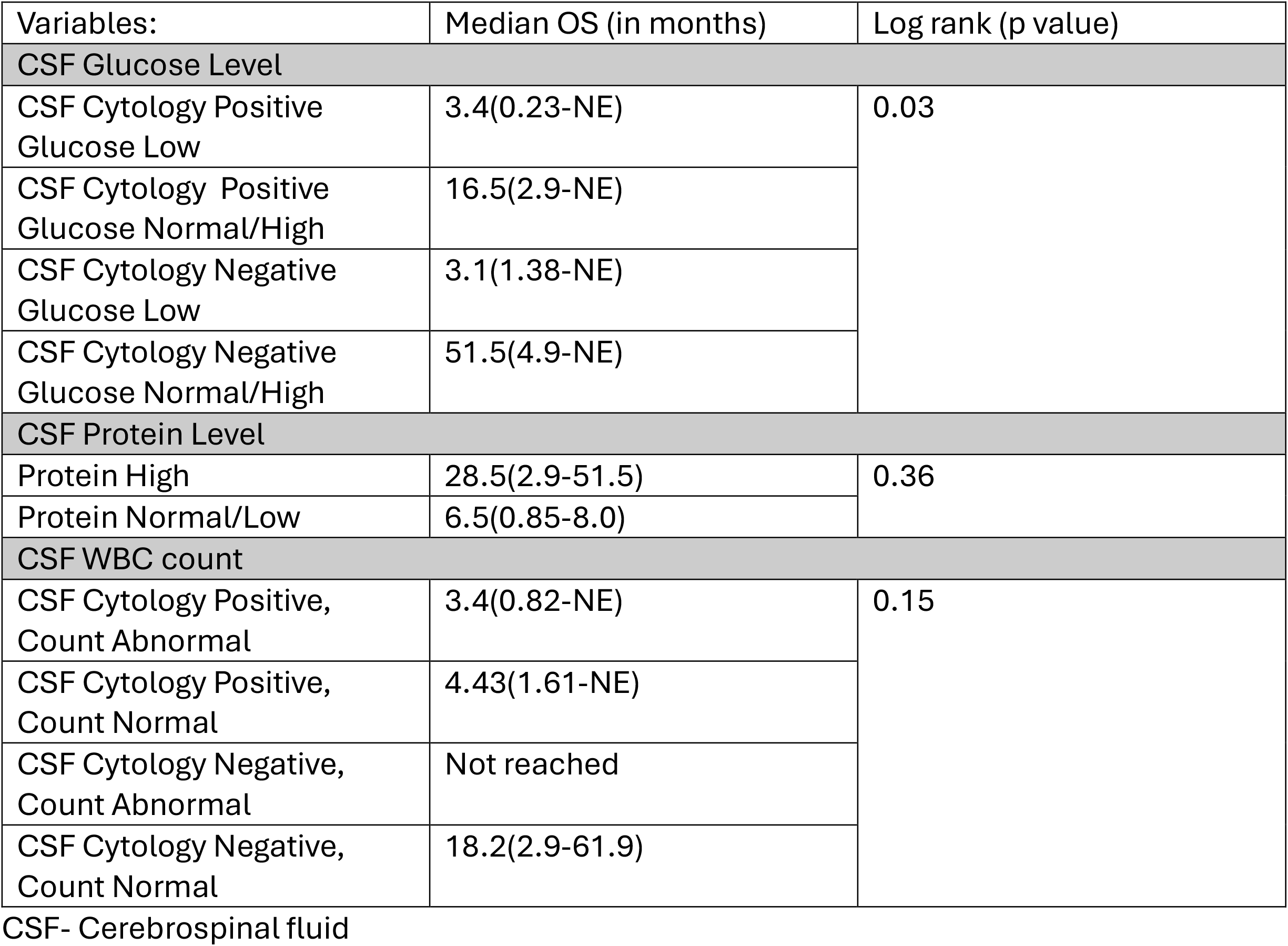
Unadjusted comparison of OS.

| Variables: | Median OS (in months) | Log rank (p value) |
| --- | --- | --- |
| CSF Glucose Level |  |  |
| CSF Cytology Positive<br>Glucose Low | 3.4(0.23-NE) | 0.03 |
| CSF Cytology Positive<br>Glucose Normal/High | 16.5(2.9-NE) |  |
| CSF Cytology Negative<br>Glucose Low | 3.1(1.38-NE) |  |
| CSF Cytology Negative<br>Glucose Normal/High | 51.5(4.9-NE) |  |
| CSF Protein Level |  |  |
| Protein High | 28.5(2.9-51.5) | 0.36 |
| Protein Normal/Low | 6.5(0.85-8.0) |  |
| CSF WBC count |  |  |
| CSF Cytology Positive,<br>Count Abnormal | 3.4(0.82-NE) | 0.15 |
| CSF Cytology Positive,<br>Count Normal | 4.43(1.61-NE) |  |
| CSF Cytology Negative,<br>Count Abnormal | Not reached |  |
| CSF Cytology Negative,<br>Count Normal | 18.2(2.9-61.9) |  |

CSF- Cerebrospinal fluid

**Table 4.** Multivariate Analyses.

| Variables | Hazard ratio | 95% CI | P value |
| --- | --- | --- | --- |
| **CSF glucose** |  |  |  |
| CSF glucose Low (vs. Normal/High) | 4.64 | (1.71, 13.2) | 0.003 |
| Interval from metastatic disease to LMD diagnosis (> vs <= 12 months) | 2.85 | (1.14, 7.82) | 0.02 |
| **CSF protein** |  |  |  |
| CSF protein High(vs Normal/Low) | 0.54 | (0.20, 1.58) | 0.24 |
| Interval from metastatic disease to LMD diagnosis > 12 months (vs <= 12 months) | 2.51` | (1.04, 6.68) | 0.04 |
| CSF cytology Positive (vs Negative) | 2.75 | (1.10, 7.20) | 0.03 |
| **CSF WBC count** |  |  |  |
| CSF WBC count Abnormal vs Normal | 1.86 | (0.73, 4.66) | 0.19 |
| Interval from metastatic disease to LMD diagnosis (> vs <= 12 months) | 2.67 | (1.05, 7.50) | 0.04 |

**Figure 2.**
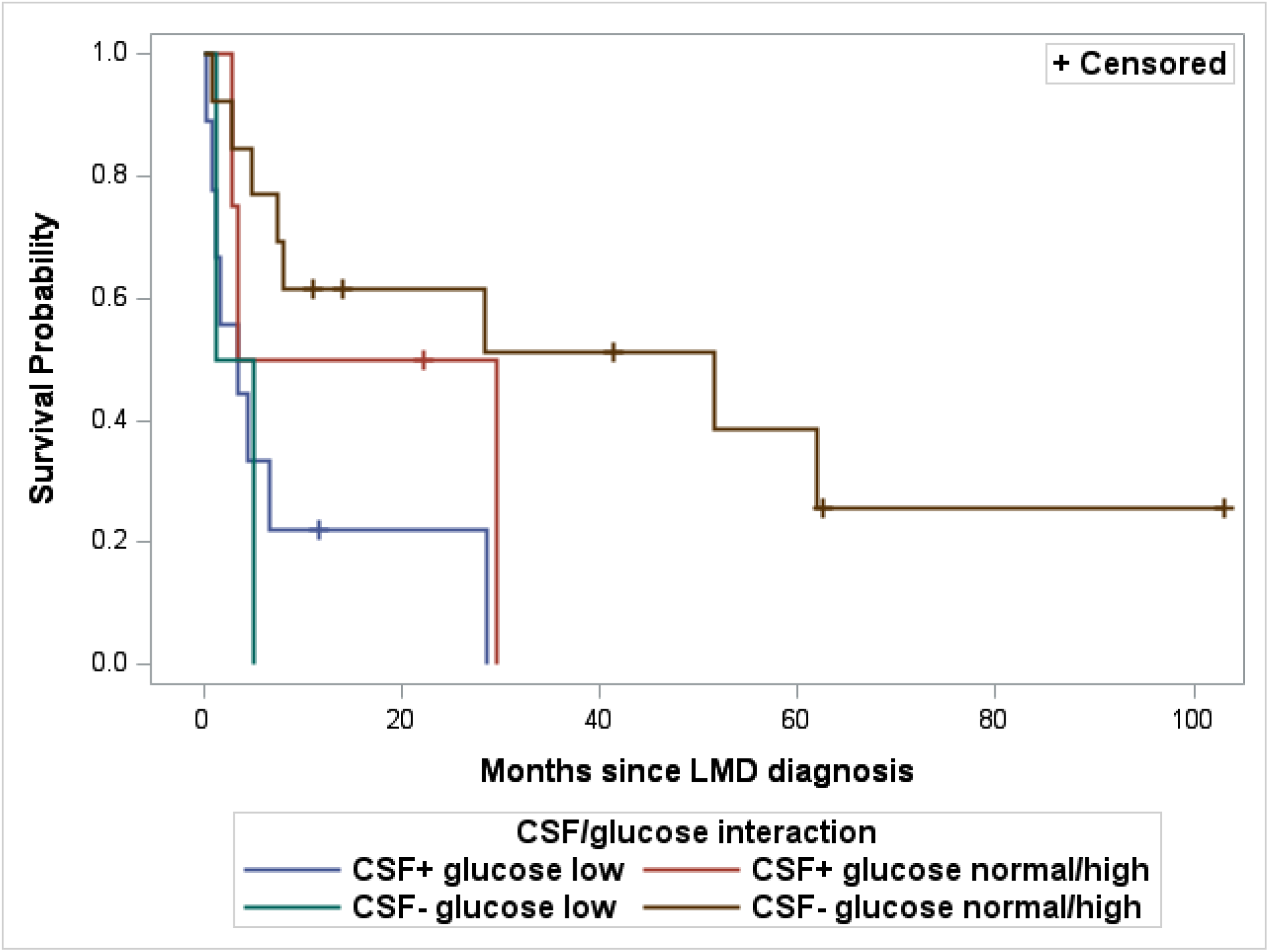
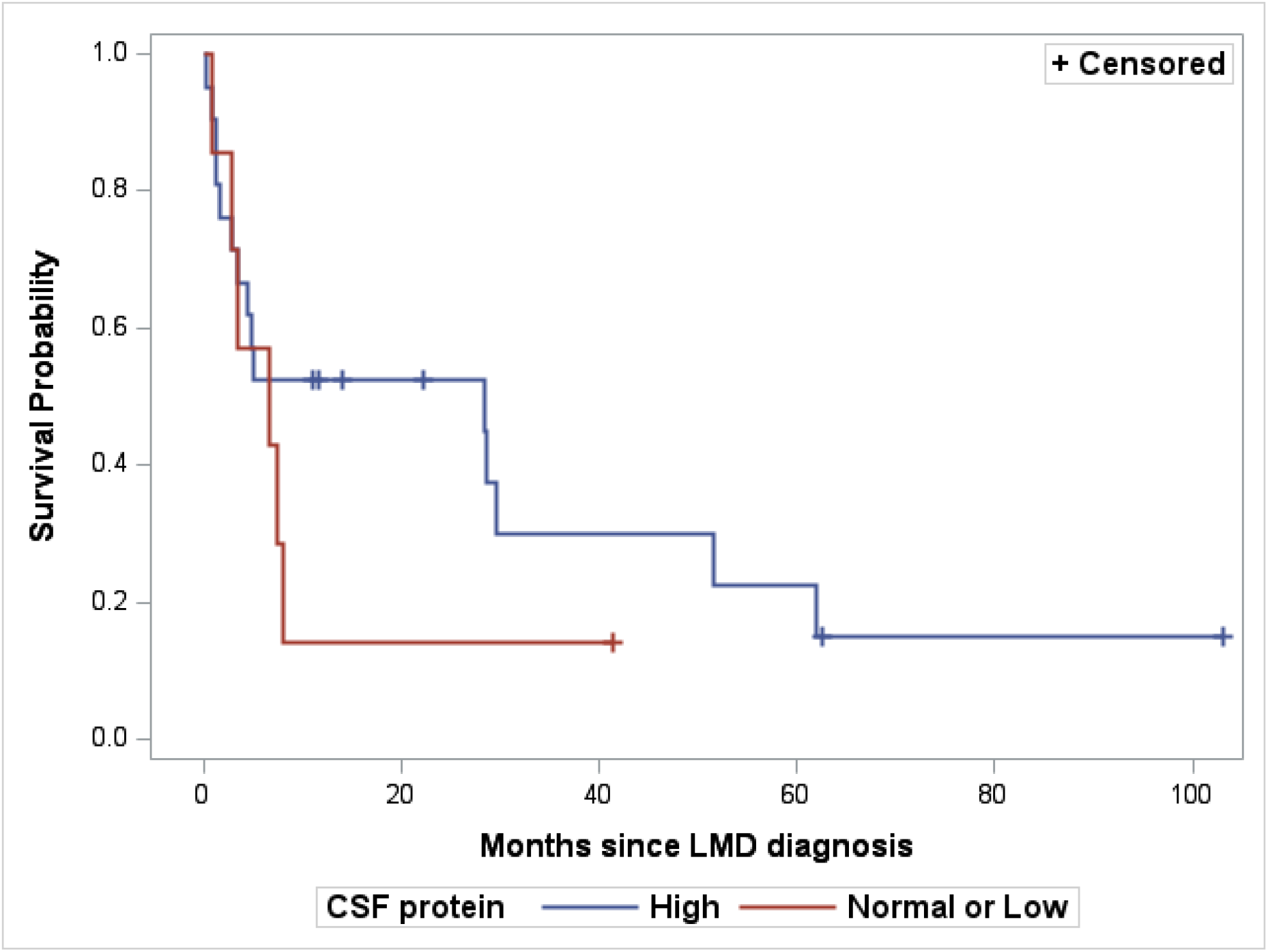
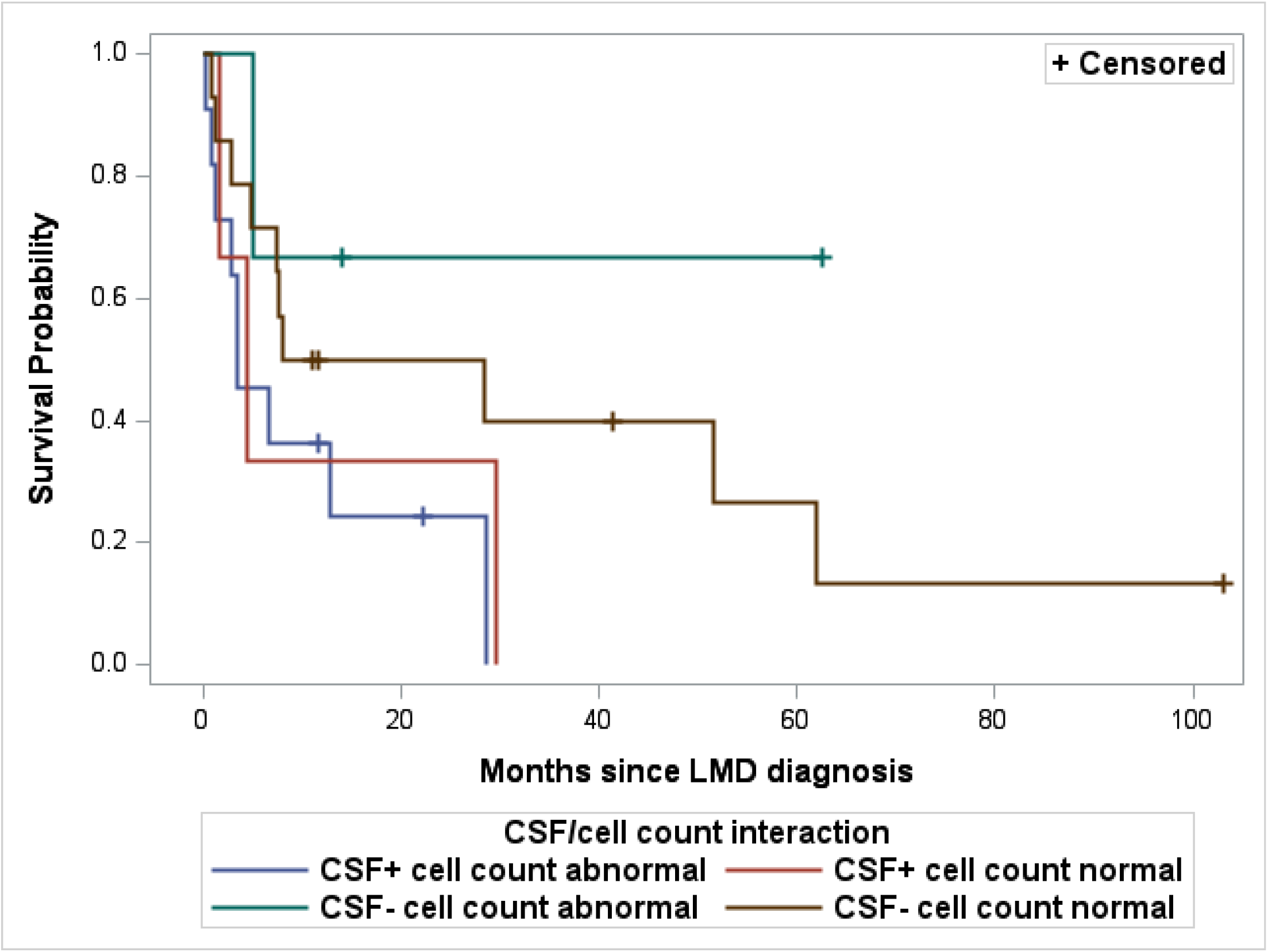
Kaplan Meier Overall Survival Analysis: A. Stratified by CSF Glucose (Low vs. Normal or High) and CSF Cytology (+ vs. -) B. Stratified by CSF Protein (High vs. Normal or Low) C. Stratified by CSF WBC Count (Normal vs. Abnormal) and CSF Cytology (+ vs. -)

*Survival based on CSF Protein:* Both in unadjusted analyses and multivariate analysis, there was insufficient evidence of association between CSF protein level and OS (Tables 3,4; Figure 2b).

*Survival Based on CSF WBC count:* Both in unadjusted analyses and in multivariate analysis, there was insufficient evidence of association between CSF WBC count and OS (Tables 3,4; Figure 2c).

## Discussion

Few studies have explored the prognostic significance of CSF biomarkers-glucose and protein among patients with breast cancer LMD, yielding inconsistent results. Griguolo et al. investigated 153 breast cancer patients with LMD retrospectively. The study showed that breast cancer LMD patients with CSF glucose of <3 mmol/L had a median OS of 2.8 (1.5-4.1) months compared to 7.4 (4.7-10.0) months [HR 1.84 (1.18-2.88) p=0.007] in those with CSF glucose >=3 mmol/L. Similarly, the cohort with CSF protein >= 1 g/L had OS of 2.4 months (0.6-4.3) compared to 7.4 months (4.1-10.7) among the cohort with CSF protein < 1 g/L [HR 2.15 (1.39-3.34), p=0.001]. Hence, this study demonstrated a significant association of low CSF glucose and high CSF protein, separately associated with worse survival among breast cancer LMD patients ^5^ Palma et al. studied 50 patients with LMD from different primary malignancies (35% breast cancer). This retrospective study failed to show significant prognostic significance of CSF glucose levels. In contrast, patients with high CSF protein levels had worse survival than those with low CSF protein levels [HR 0.39 (0.1-0.51), p=0.003].^6^ Clatot et al. investigated 24 patients with breast cancer LMD retrospectively, all of whom received intrathecal methotrexate. This study showed that CSF glucose and protein levels were not statistically associated with survival.^7^ Boogerd et al. studied 58 breast cancer patients with LMD retrospectively, the majority of whom were treated with intrathecal chemotherapy.^3^ In this study, in multivariate analyses, low CSF glucose and high CSF proteins were associated with worse survival. In a large retrospective study enrolling 512 LMD patients from different primary malignancies (including 96 patients with breast cancer), multivariate analyses showed low CSF protein was significantly associated with better survival. CSF glucose was not assessed in this study.^8^ Similarly, Bruna et al. studied LMD cases from different primary malignancies. They included 30 cases of breast cancer LMD and found that a high CSF glucose was associated with a better prognosis.^9^ Our study showed that low CSF glucose was significantly associated with poor survival compared to the group with normal or high CSF glucose. However, our study did not show a significant association between CSF protein levels and OS. These inconsistencies across studies could be due to the retrospective nature of all studies, slightly different study populations (e.g., a couple of studies included only breast cancer LMD treated with intrathecal therapies, while others included any breast cancer LMD), and different sample sizes. In addition, different studies have different cut-offs to define normal vs abnormal levels of these CSF parameters. In our literature search, we didn’t find any prognostic data on CSF WBC count. Our study could not show any significant association between CSF WBC count and OS.

A prior study from this data set suggested that breast cancer LMD patients with a positive CSF cytology had worse survival compared to patients with a negative CSF cytology.^4^ However, in this study, we found a significant association between CSF cytology results and CSF glucose levels. In multivariate analysis, CSF glucose level was significantly associated with OS; however, CSF cytology result was not. A larger study is needed to confirm this finding.

## Conclusion

This small single-institute retrospective study showed that low CSF glucose is associated with worse overall survival among breast cancer patients with leptomeningeal disease. There was insufficient evidence to suggest that CSF protein level or CSF WBC counts were associated with survival in this study. A larger prospective study is required to confirm these findings.

## Data Availability

All data produced in the present study are available upon reasonable request to the authors.

